# Validation of Real-World Case Definitions for COVID-19 Diagnosis and Severe COVID-19 Illness Among Patients Infected with SARS-CoV-2: Translation of Clinical Trial Definitions to Real-World Settings

**DOI:** 10.1101/2023.09.12.23295441

**Authors:** Mei Sheng Duh, Catherine Nguyen, Heather Rubino, Christopher Herrick, Rose Chang, Maral DerSarkissian, Yichuan Grace Hsieh, Azeem Banatwala, Louise H. Yu, Gregory Belsky, Marykate E. Murphy, Janet Boyle-Kelly, Andrew Cagan, Bruce E. Stangle, Pierre Y. Cremieux, Francesca Kolitsopoulos, Shawn N. Murphy

## Abstract

**Purpose:** This study assessed the performance of International Classification of Diseases 10th Revision, Clinical Modification (ICD-10-CM) coronavirus disease 2019 (COVID-19) diagnostic code U07.1 against polymerase chain reaction (PCR) test results (Objective 1), and electronic medical record (EMR)-based codified algorithm for severe COVID-19 illness based on endpoints used in the Pfizer-BioNTech COVID-19 vaccine trial against chart review (Objective 2).

**Methods:** This retrospective, longitudinal cohort study used EMR data from the Mass General Brigham COVID-19 Data Mart (3/1/2020–11/19/2020) for adult patients with ≥1 PCR test, antigen test, or code U07.1 (Objective 1) and adult patients with a positive PCR test hospitalized with COVID-19 (Objective 2).

**Results:** Among 354,124 patients in Objective 1, 96% had ≥1 PCR test (including 6% with ≥1 positive PCR test; 11% with ≥1 code U07.1). Code U07.1 had low sensitivity (54%) and positive predictive value (PPV; 63%) but high specificity (97%) against the PCR test. Among 300 patients hospitalized for COVID-19 randomly sampled for chart review in Objective 2, the EMR-based case definition for severe COVID-19 illness had high PPV (>95%), showing better performance than severe/critical COVID-19 endpoints defined by the World Health Organization (PPV: 79%).

**Conclusions:** COVID-19 diagnosis based on ICD-10-CM code U07.1 had inadequate sensitivity and requires confirmation by PCR testing. The EMR-based case definition showed high PPV and can be used to identify cases of severe COVID-19 illness in real-world datasets. These findings highlight the importance of validating outcomes in real-world data, and can guide researchers analyzing COVID-19 data when PCR tests are not readily available.

**KEY POINTS:**

- This study evaluated the performance of International Classification of Diseases 10th Revision, Clinical Modification (ICD-10-CM) codes and an electronic medical record (EMR)-based algorithm for identifying coronavirus disease 2019 (COVID-19) diagnosis and severe COVID-19 illness in real-world data.
- ICD-10-CM code U07.1 for COVID-19 had low sensitivity and positive predictive value (PPV) against PCR tests.
- The EMR-based algorithm for severe COVID-19 illness developed from the Pfizer– BioNTech COVID-19 vaccine trial had high PPV against chart review, and may be used to identify severe cases in real-world data.
- These results highlight the importance of validating outcomes when conducting analyses of real-world datasets.

**PLAIN LANGUAGE SUMMARY:** As polymerase chain reaction (PCR) tests for coronavirus disease 2019 (COVID-19) diagnosis are becoming less frequently used and there is no standard definition of severe COVID-19 illness, it is important to have a way of correctly identifying COVID-19 diagnosis or severe COVID-19 illness in real-world data (e.g., electronic medical records [EMRs]). This study examined: 1) how a diagnosis code for COVID-19 used in EMRs (i.e., U07.1) compares to PCR test results in terms of accurately identifying patients with COVID-19; and 2) whether a definition for severe COVID-19 illness developed based on the Pfizer–BioNTech COVID-19 vaccine trial and a definition used by the World Health Organization [WHO]) can be used to accurately identify patients with severe COVID-19 illness in EMRs. The results showed that code U07.1 was not very accurate in identifying patients with COVID-19. On the other hand, the developed definition for severe COVID-19 illness was more accurate than the WHO definition and was able to identify most patients with severe COVID-19 illness in real-world data.

## INTRODUCTION

Several vaccines have been developed to respond to the coronavirus disease 2019 (COVID-19) pandemic following the emergence of severe acute respiratory syndrome coronavirus 2 (SARS-CoV-2), which was identified in the United States (US) in March 2020.^1^ For a COVID-19 vaccine to be approved by the US Food and Drug Administration (FDA), it must demonstrate ≥50% efficacy compared with placebo in preventing COVID-19 illness.^2,3^ In the clinical trial for the SARS-CoV-2 RNA vaccine BNT162b2 developed by Pfizer in collaboration with BioNTech (NCT04368728), the vaccine showed 95% efficacy against COVID-19 and 90% efficacy against severe COVID-19 illness, and was well tolerated.^4,5^ Based on these results, BNT162b2 was granted emergency use authorization (EUA) on December 11, 2020^6^ and full approval on August 23, 2021.^7^

Despite the availability of vaccines, only about two-thirds (67.5%) of the US population was fully vaccinated as of September 6, 2022.^8^ As new SARS-CoV-2 variants have emerged, unvaccinated persons have shown consistently higher rates of hospitalization and death from COVID-19 than those who are vaccinated.^9,10^ Early identification and treatment of severe cases can prevent clinical progression^11^ but remain challenging. Reverse transcription polymerase chain reaction (PCR) tests are considered the gold standard diagnostic method^12^ but are resource-intensive and therefore not ideal for point-of-care testing. Moreover, there is no consistent definition of COVID-19 illness severity. The development and validation of COVID-19 diagnostic measures and severe illness endpoints used outside of clinical trials that can accurately identify these measures in varied sources such as electronic medical records (EMRs) and claims databases are important for assessing the efficacy and safety of COVID-19 vaccines and treatments in real-world settings.

As the performance of the International Classification of Diseases 10th Revision, Clinical Modification (ICD-10-CM) diagnosis code U07.1 may vary over time and by population, the performance of the code was validated against the gold-standard PCR test (Part 1). Additionally, a case definition of severe COVID-19 illness in patients hospitalized with COVID-19 using EMR data was evaluated by medical chart review (Part 2).

## METHODS

### Study design

A retrospective, longitudinal cohort study was conducted to evaluate COVID-19 diagnostic measures. The index date was defined as the date of the earliest COVID-19 diagnostic measure (i.e., PCR molecular test, diagnosis code U07.1, or antigen test) (Part 1). To assess the performance of an EMR-based case definition for severe COVID-19 illness, the index date was defined as the date of hospitalization with COVID-19 (readmission within 3 days was considered a continuation of the previous hospitalization), and the baseline period was defined as the 3-month period preceding the index date (Part 2). The observation period in both parts of the study was the time from the index date to the earliest of death, loss to follow-up, or end of data availability.

### Data source and extraction

The Mass General Brigham (MGB) Research Patient Data Registry COVID-19 Data Mart provided deidentified inpatient and outpatient EMR data from the MGB healthcare system for more than 6.5 million patients with over 2 billion records. The MGB system consists of 8 major hospitals affiliated with Harvard Medical School in Massachusetts (Massachusetts General Hospital, Brigham and Women’s Hospital, Brigham and Women’s Faulkner Hospital, Massachusetts Eye and Ear Hospital, McLean Hospital, Newton-Wellesley Hospital, North Shore Medical Center, and Spaulding Rehabilitation Hospital).^13^ Data elements include demographics, providers, visits, diagnoses, medications, procedures, laboratory, microbiology, and work-up reports (e.g., discharge, operative, radiology). Codified EMR data from March 1, 2020 (when PCR test results were first available) to November 19, 2020 were extracted for this study.

### Study population

Part 1 of the study included patients aged 18–85 years with ≥1 diagnostic measure of COVID-19 at the time the measure was recorded. All patients were included in the calculation of frequency of diagnostic measures, but only those with PCR test results and diagnosis code U07.1 were included in the validation analysis. Part 2 included patients aged 18–85 years with a positive PCR test and hospitalization associated with COVID-19 (defined as ≥1 positive PCR test in the 14 days prior to hospital admission or ≥1 positive PCR test in the 2 days following hospital admission).

### Severe and critical COVID-19 illness endpoint definitions

Severe COVID-19 illness endpoints were defined using an algorithm derived from the Pfizer-BioNTech COVID-19 vaccine trial^5^ that included diagnosis codes, procedure codes, laboratory test results, and medication records. Endpoints used by the WHO^14^ to classify COVID-19 illness as severe or critical were also similarly identified. The EMR-based case definition was validated through medical chart review in a randomly selected sample of 300 patients with and 100 patients without severe COVID-19 illness in MGB’s codified data according to the Pfizer-BioNTech COVID-19 vaccine trial definition. Severe or critical COVID-19 illness based on the WHO definition was also validated through medical chart review for the subsample of patients who met both the vaccine trial and WHO definitions of severe COVID-19 illness.

### Statistical analysis

Data were analyzed using SQL Server Management Studio 18 (Microsoft, Redmond, WA, USA) and SAS Enterprise Guide v7.1 (SAS Institute, Cary, NC, USA).

For Part 1, the frequency of PCR tests and diagnosis code U07.1 was assessed over the whole study period and by month. Characteristics were summarized with frequency distributions for categorical variables and mean (standard deviation [SD]) and median for continuous variables. The closest record of a diagnosis code within 2 days of a PCR test result was validated against the latter. If a patient had multiple PCR tests, the first one during the period of interest was used. Patients were classified as true positive (TP) if they had code U07.1 and were positive by PCR; false positive (FP) if they had code U07.1 but were negative by PCR; true negative (TN) if they lacked code U07.1 and were negative by PCR; and false negative (FN) if they lacked code U07.1 but were positive by PCR. Sensitivity was calculated as TP/(TP+FN); specificity as TN/(TN+FP); positive predictive value (PPV) as TP/(TP+FP); and negative predictive value (NPV) as TN/(TN+FN).

For Part 2, baseline characteristics of patients hospitalized with COVID-19 were summarized. To validate the EMR-based case definition, 300 patients with severe COVID-19 illness were classified as TP or FP and 100 patients without severe illness were classified as TN or FN based on chart review (the gold standard), and PPV and NPV were calculated.

## RESULTS

In Part 1, 354,124 patients met the inclusion criteria and had ≥1 COVID-19 diagnostic measure from March 1 to November 19, 2020. Part 2 of this study analyzed 3,580 of these patients who had ≥1 positive PCR test and were hospitalized with COVID-19 (**Fig. S1**).

### Distribution of COVID-19 diagnostic measures

Overall, 94.6% of patients had ≥1 PCR test and 6.5% had ≥1 positive PCR test (**Table S1**). Of the 726,652 PCR tests administered, 5.9% were positive. From March to November 2020, 11.2% of patients had ≥1 diagnosis code U0.71; the mean (SD) number of codes per patient was 3.0 (5.4).

### Trends in COVID-19 diagnostic tests and coding

The number of patients per month receiving a PCR test increased from 11,412 in March to 71,637 in August and remained elevated through November (**Fig. 1a**). The number of patients per month with diagnosis code U07.1 increased from 906 in March to 10,700 in April but declined between May (9,779) and June (5,036), remaining stable thereafter.

**Figure 1.**
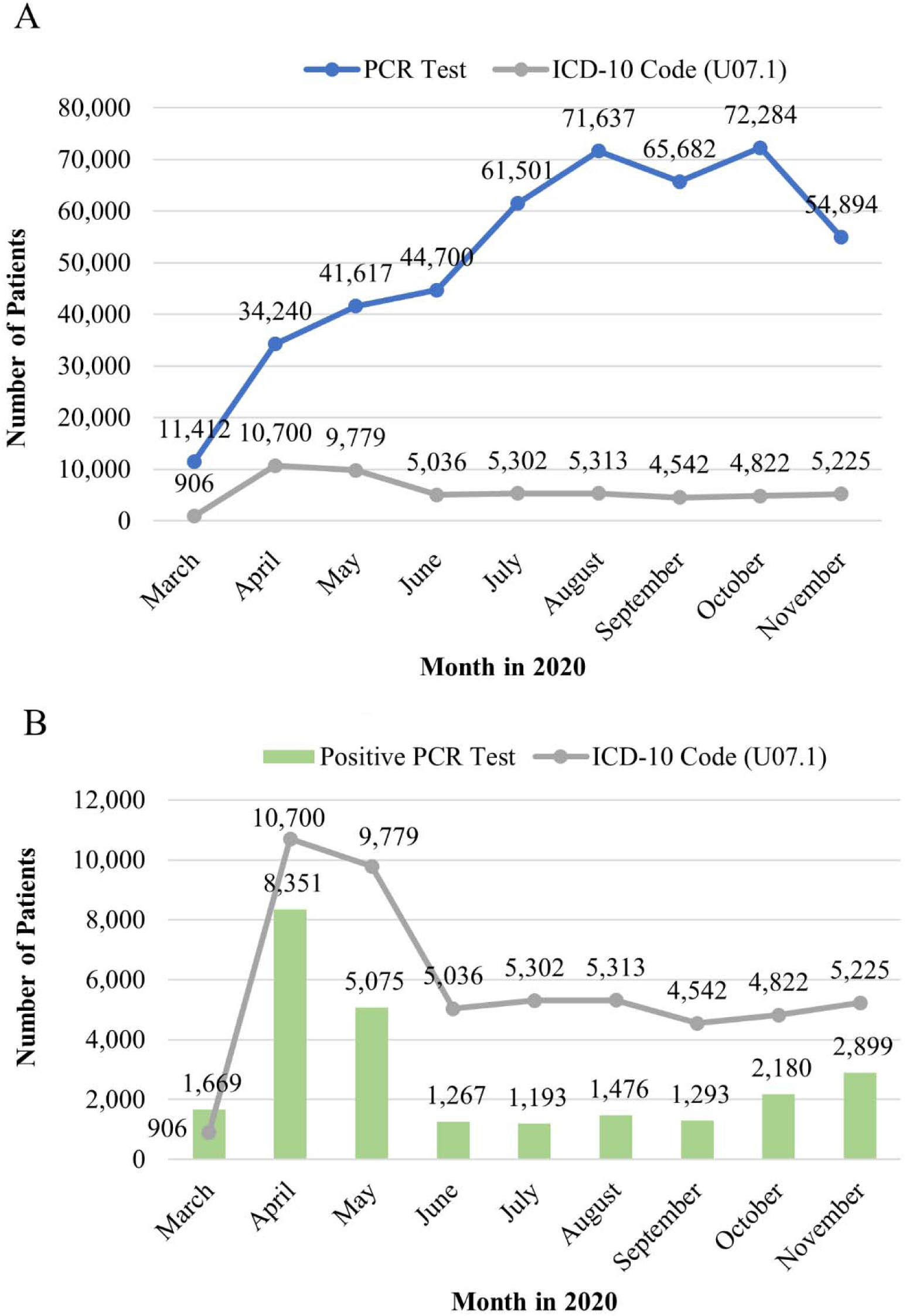
Number of patients with each COVID-19 diagnostic measure and positive PCR test or ICD-10-CM code by month. **A.** Number of patients with PCR test or ICD-10-CM code by month. **B.** Number of patients with positive COVID-19 PCR test or ICD-10-CM code by month. Data for November were available only up to November 19, 2020.

Positive PCR tests and diagnosis code U07.1 peaked in April before declining in May (**Fig. 1b**) and remained relatively stable from June through November. There were consistently more patients with diagnosis code U07.1 than with a positive PCR test from April to November.

### Validation of the diagnosis code against the PCR test

The sensitivity of diagnosis code U07.1 against the PCR test was 53.8% from March to November and varied over the study period, being highest in April to June (≥60%) and lowest in October and November (∼20%) (**Table 1**). The specificity of the diagnosis code remained high (>90%) during the study period and was 96.6% overall. The PPV of the diagnosis code was 47.4% over the study period and was highest (>70%) in March and April. The NPV of the diagnosis code was high (>88%) across all months and was 97.3% overall.

**Table 1.**
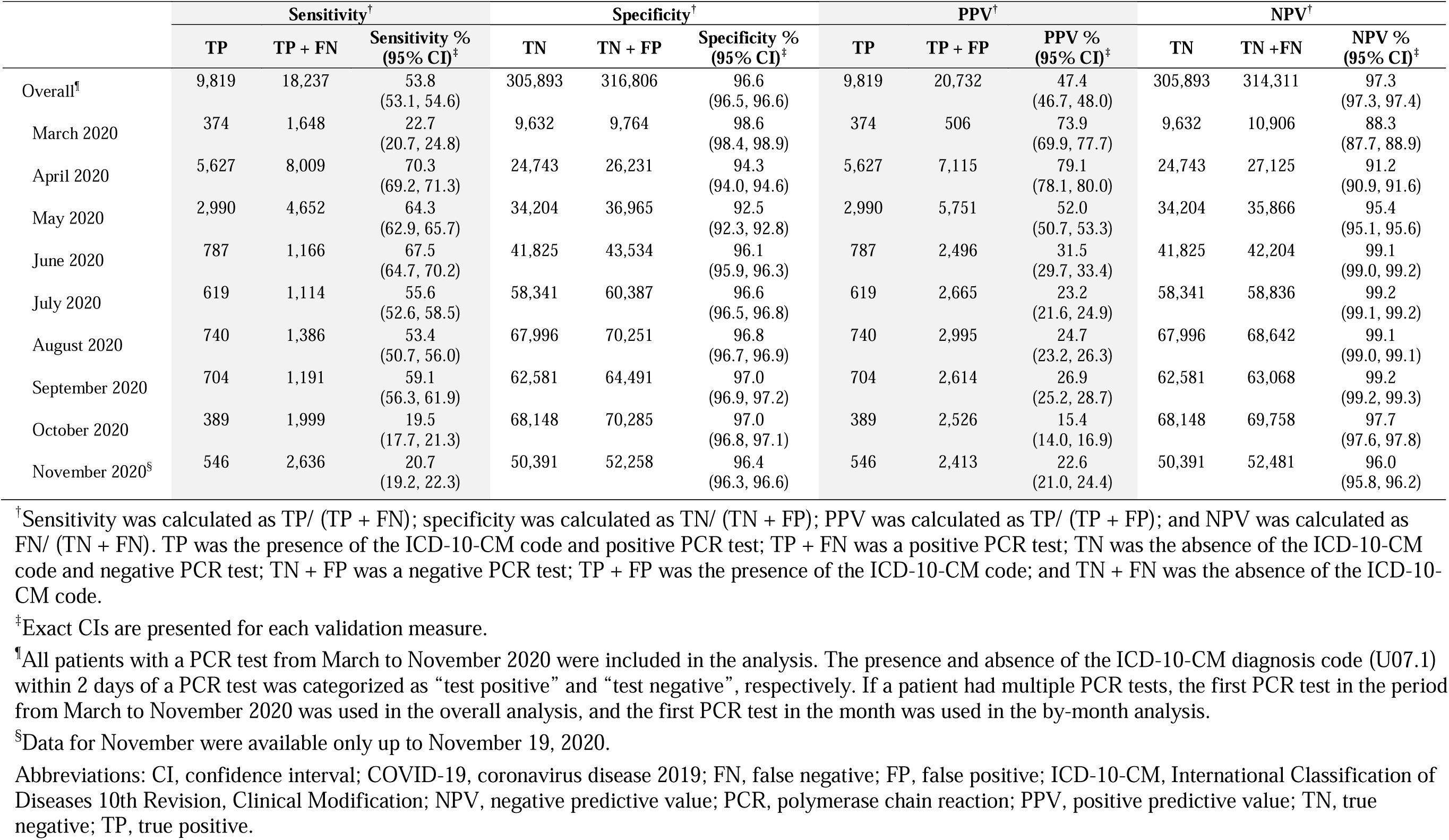
Validation of COVID-19 ICD-10-CM diagnosis code against PCR test.

### Characteristics of the population with severe COVID-19 illness

On the index date, patients hospitalized with COVID-19 had a mean age of 59 years; 57% were male, and a large proportion (42%) were overweight or obese (body mass index ≥25 kg/m^2^) (**Table 2**). Common comorbidities included hypertension (44%), chronic kidney disease/dialysis (31%), and diabetes mellitus (30%). The first positive PCR test result for most patients (64%) was on the index date and was performed in an inpatient setting (57%). The main symptoms experienced by patients were shortness of breath (33%), cough (26%), and fever (25%).

**Table 2.** Baseline characteristics of patients hospitalized with COVID-19.

| Characteristic<sup>†</sup> | All patients<br>hospitalized with<br>COVID-19<br>N = 3,580 |
| --- | --- |
| **Demographic characteristics** |  |
| Age on index date, years |  |
| Mean ± SD | 59.0 ± 16.3 |
| Median (IQR) | 60.0 (48.0, 72.0) |
| Age on index date, years, n (%) |  |
| 18–29 | 177 (4.9) |
| 30–39 | 351 (9.8) |
| 40–49 | 450 (12.6) |
| 50–59 | 711 (19.9) |
| 60–69 | 799 (22.3) |
| 70–79 | 717 (20.0) |
| 80–86 | 375 (10.5) |
| Sex, n (%) |  |
| Male | 2,027 (56.6) |
| Female | 1,553 (43.4) |
| Race, n (%) |  |
| White | 1,704 (47.6) |
| Black or African American | 631 (17.6) |
| Asian | 116 (3.2) |
| American Indian/Alaska Native | 9 (0.3) |
| Native Hawaiian/Pacific Island | 7 (0.2) |
| Other<sup>‡</sup> | 896 (25.0) |
| Unknown | 217 (6.1) |
| Smoking status, n (%)<sup>¶</sup> |  |
| Never smoker | 1,418 (39.6) |
| Former smoker | 372 (10.4) |
| Current smoker | 293 (8.2) |
| Unknown | 1,497 (41.8) |
| **Clinical characteristics** |  |
| BMI, kg/m<sup>2</sup>, n (%)<sup>¶</sup> |  |
| Underweight (<18.5) | 31 (0.9) |
| Normal weight (18.5–24.9) | 353 (9.9) |
| Overweight (25–29.9) | 594 (16.6) |
| Obese (≥30) | 909 (25.4) |
| Unknown | 1,693 (47.3) |
| Quan-CCI<sup>§</sup> |  |
| Mean ± SD | 1.3 ± 1.9 |
| Median (IQR) | 0.0 (0.0, 2.0) |
| Comorbidities, n (%)<sup>□</sup> |  |
| Hypertension | 1,575 (44.0) |
| Chronic kidney disease/dialysis | 1,099 (30.7) |
| Diabetes mellitus | 1,064 (29.7) |
| Hyperlipidemia | 985 (27.5) |
| Chronic obstructive pulmonary disease/interstitial lung disease | 793 (22.2) |
| Coronary artery disease | 655 (18.3) |
| Asthma | 333 (9.3) |
| Bleeding diathesis or condition associated with prolonged bleeding | 324 (9.1) |
| Cancer | 306 (8.5) |
| Liver disease | 260 (7.3) |
| Venous thromboembolism | 260 (7.3) |
| Autoimmune disease | 195 (5.4) |
| HIV/HBV/HCV | 108 (3.0) |
| Other immune deficiencies | 103 (2.9) |
| Solid organ transplant | 4 (0.1) |
| Concomitant medications, n (%)<sup>#</sup> |  |
| Antibiotics | 1,925 (53.8) |
| NSAIDs | 1,114 (31.1) |
| Statins | 1,192 (33.3) |
| Corticosteroids | 729 (20.4) |
| Previous vaccine anaphylaxis, n (%)<sup>□</sup> | 4 (0.1) |
| **Disease characteristics** |  |
| Vital signs on index date, n (%)<sup>††</sup> |  |
| Patients with temperature available | 3,556 (99.3) |
| <37.5°C (normal) | 1,705 (47.9) |
| 37.5°C–38.0°C | 639 (18.0) |
| 38.1°C–39.0°C | 807 (22.7) |
| >39.0°C | 405 (11.4) |
| Patients with heart rate available | 3,567 (99.6) |
| <125 beats/min (normal) | 3,147 (88.2) |
| ≥125 beats/min | 420 (11.8) |
| Patients with respiratory frequency available | 3,515 (98.2) |
| <30 breaths/min (normal) | 2,402 (68.3) |
| ≥30 breaths/min | 1,113 (31.7) |
| Patients with SpO<sub>2</sub> available | 3,564 (99.6) |
| ≤93% | 2,003 (56.2) |
| >93% (normal) | 1,561 (43.8) |
| Patients with SBP available | 3,559 (99.4) |
| <90 mm Hg | 376 (10.6) |
| ≥90 mm Hg | 3,183 (89.4) |
| Time from first positive PCR test to index date, n (%) |  |
| 7–14 days prior to index date | 222 (6.2) |
| ≤7 days prior to index date | 640 (17.9) |
| On index date | 2,291 (64.0) |
| ≤2 days after index date | 427 (11.9) |
| Care setting at COVID-19 diagnosis (i.e., positive PCR test), n (%)<sup>‡‡</sup> |  |
| Inpatient | 2,036 (56.9) |
| Emergency room | 191 (5.3) |
| Outpatient | 118 (3.3) |
| Unknown | 1,235 (34.5) |
| Symptoms reported on index date, n (%)<sup>¶¶</sup> |  |
| Shortness of breath | 1,204 (33.6) |
| Cough | 932 (26.0) |
| Fever | 889 (24.8) |
| Diarrhea | 236 (6.6) |
| Myalgia | 127 (3.5) |
| Headache | 141 (3.9) |
| Sore throat | 64 (1.8) |
| Rhinorrhea | 15 (0.4) |

Data are shown as n (%) unless otherwise indicated.
<sup>†</sup>Index date (period up to 3 months prior to the date of first hospitalization with COVID-19).
<sup>‡</sup>Includes “Two or more races” or “Other race”.
<sup>¶</sup>Status on or on the date closest (in the 12 months prior) to the index date.
<sup>§</sup>Assessed during baseline period and on the index date using diagnosis codes. Reference: Quan et al. (2005). Coding algorithms for defining comorbidities in ICD-9-CM and ICD-10-CM administrative data. Medical Care, 43(11): 1130–39.
<sup>□</sup> Assessed during baseline period and on the index date using diagnosis and procedure codes.
<sup>#</sup>Assessed during baseline period and on the index date.
<sup>††</sup> Assessed on the index date. If multiple test results were available, the most severe result for each test was used.
<sup>‡‡</sup>For first positive PCR test within 14 days before or 2 days after hospitalization.
<sup>¶¶</sup>Assessed on the index date using diagnosis codes. Multiple symptoms may be reported for each patient.
Abbreviations: BMI, body mass index; CCI, Charlson Comorbidity Index; COVID-19, coronavirus disease 2019; FEU, fibrinogen equivalent units; HBV, hepatitis B virus; HCV, hepatitis C virus; HIV, human immunodeficiency virus; IQR, interquartile range; NSAID, nonsteroidal anti-inflammatory drug; PCR, polymrase chain reaction; SBP, systolic blood pressure; SD, standard deviation; SpO<sub>2</sub>, oxygen saturation.

### Frequency of severe or critical COVID-19 illness endpoints in patients hospitalized with COVID-19

Among the 3,580 patients hospitalized with COVID-19, the mean (SD) duration of hospitalization was 13.6 (17.6) days (**Table 3**); 94.9% and 75.3% met the Pfizer–BioNTech COVID-19 vaccine trial and WHO definitions of severe COVID-19, respectively, whereas 40.6% met the WHO definition of critical illness (**Tables 3** and **S2**). Among 300 patients with severe COVID-19 illness per the vaccine trial definition in codified data who were randomly selected for chart review, 89.0% had SpO_2_ ≤93% and 50.7% had respiratory frequency >30 breaths/min; 59.3% had significant acute renal, hepatic, or neurologic dysfunction; 44.7% showed evidence of shock; 43.0% were admitted to the ICU; 26.7% had respiratory failure; and the rate of in-facility death was 11.7%.

**Table 3.**
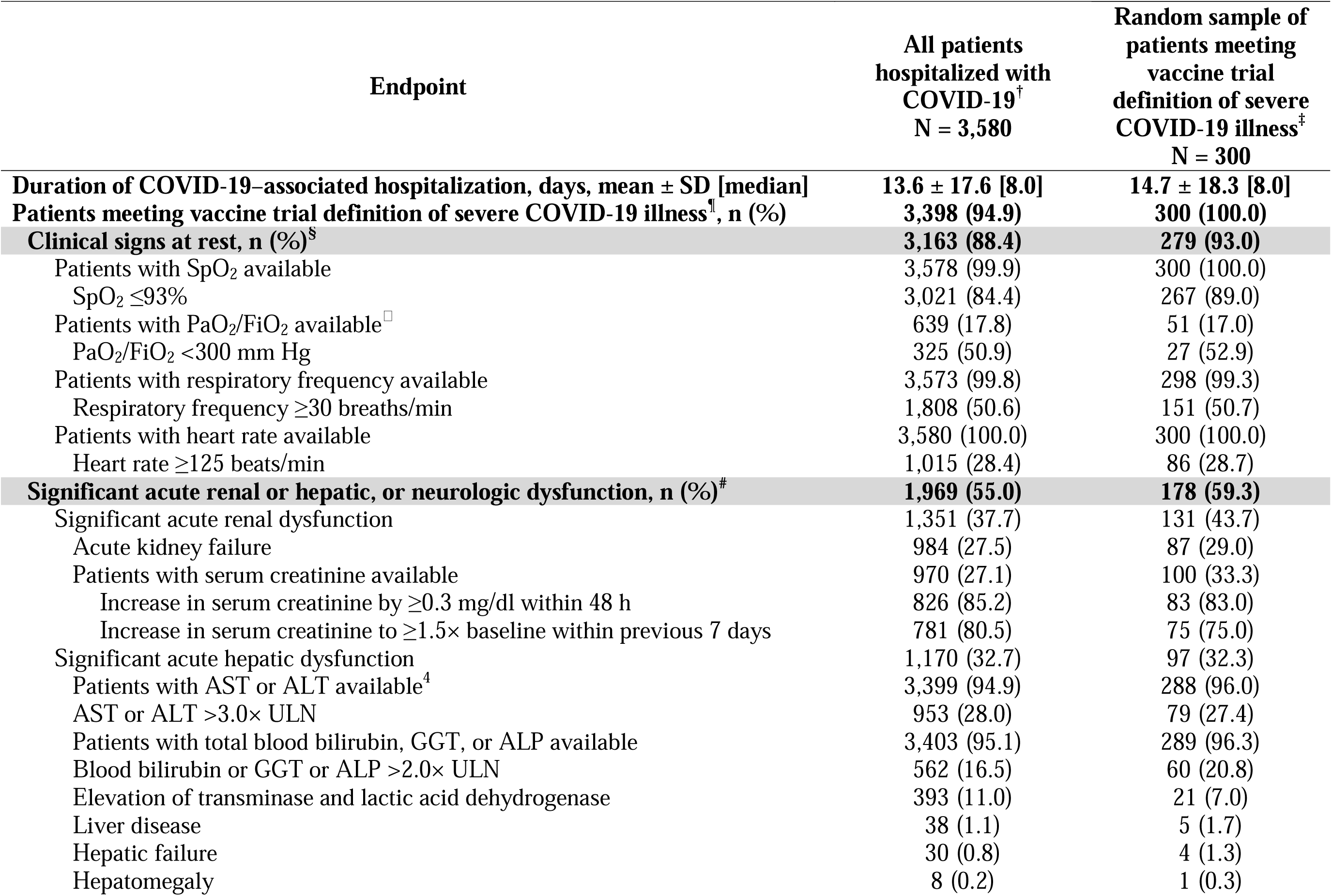
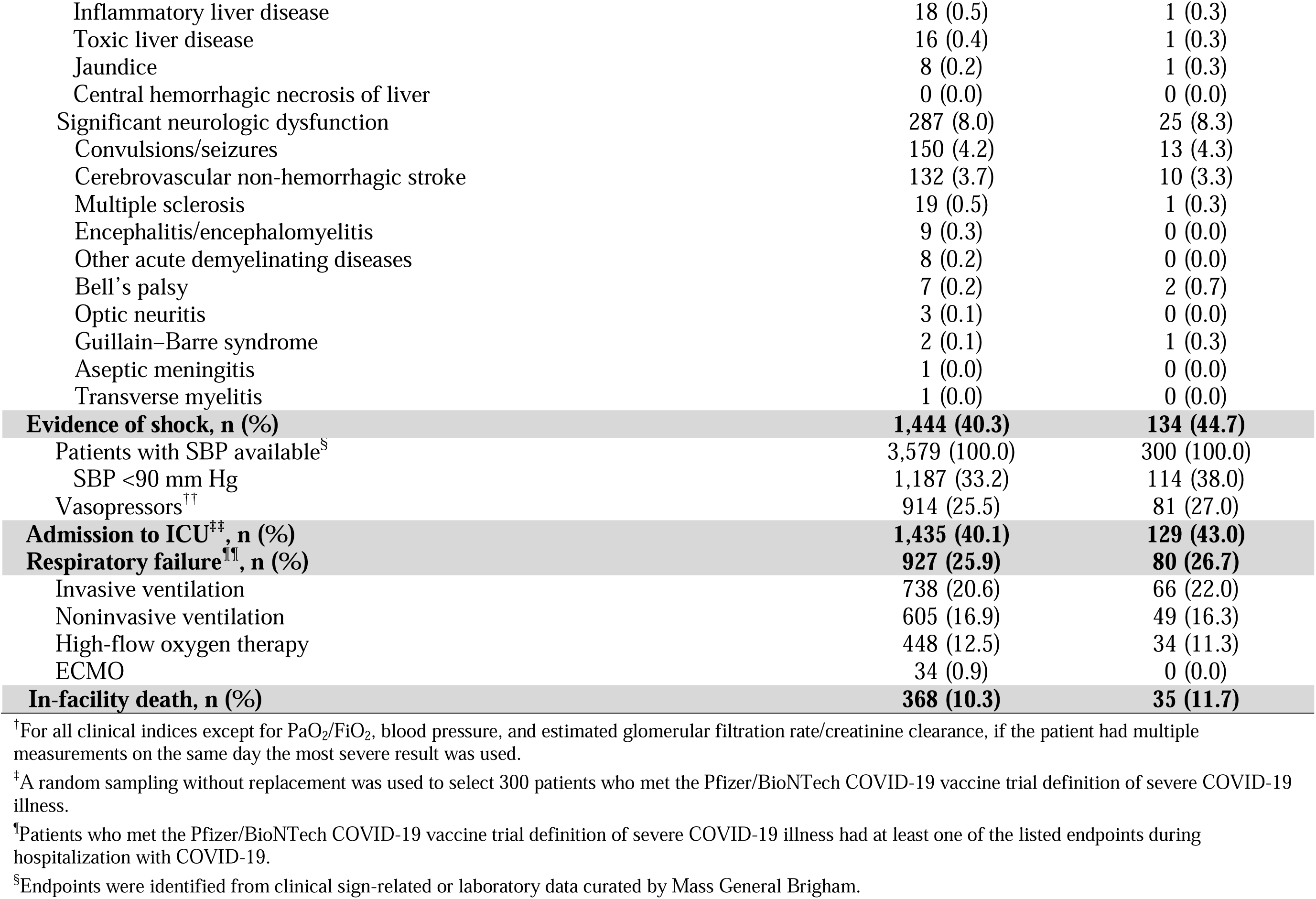
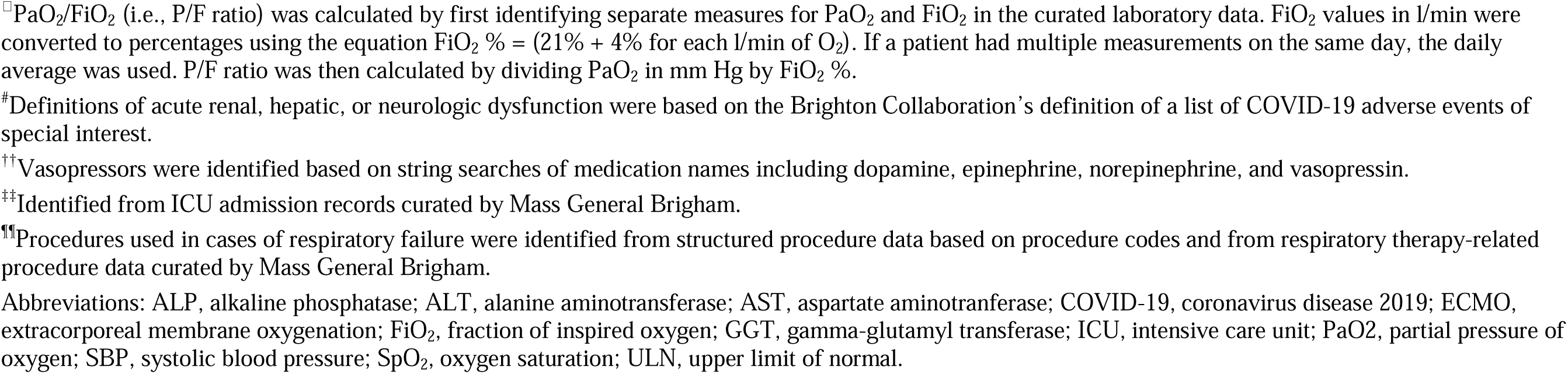
Frequency of severe COVID-19 illness endpoints in patients hospitalized with COVID-19.

Among patients meeting the WHO definition of severe COVID-19 illness,^15^ 54.0% had SpO_2_ <90% and 45.5% had a respiratory frequency >30 breaths/min, with 47.6% showing signs of severe respiratory distress (**Table S2**). Among patients meeting the WHO definition of critical COVID-19 illness, 27.8% required life-sustaining treatment including vasopressors (25.5%) and noninvasive or mechanical ventilation (37.5%).

### Validation of EMR-based case definitions for severe or critical COVID-19 illness endpoints by medical chart review

Severe COVID-19 illness endpoints in the EMR-based case definition had a high PPV (>95%), as validated through chart review (**Table 4**). The PPV of severe COVID-19 illness endpoints was high from March through June (94%–100%), but declined to <90% in subsequent months, which may have been due to the limited sample size (**Fig. S2**).

**Table 4.**
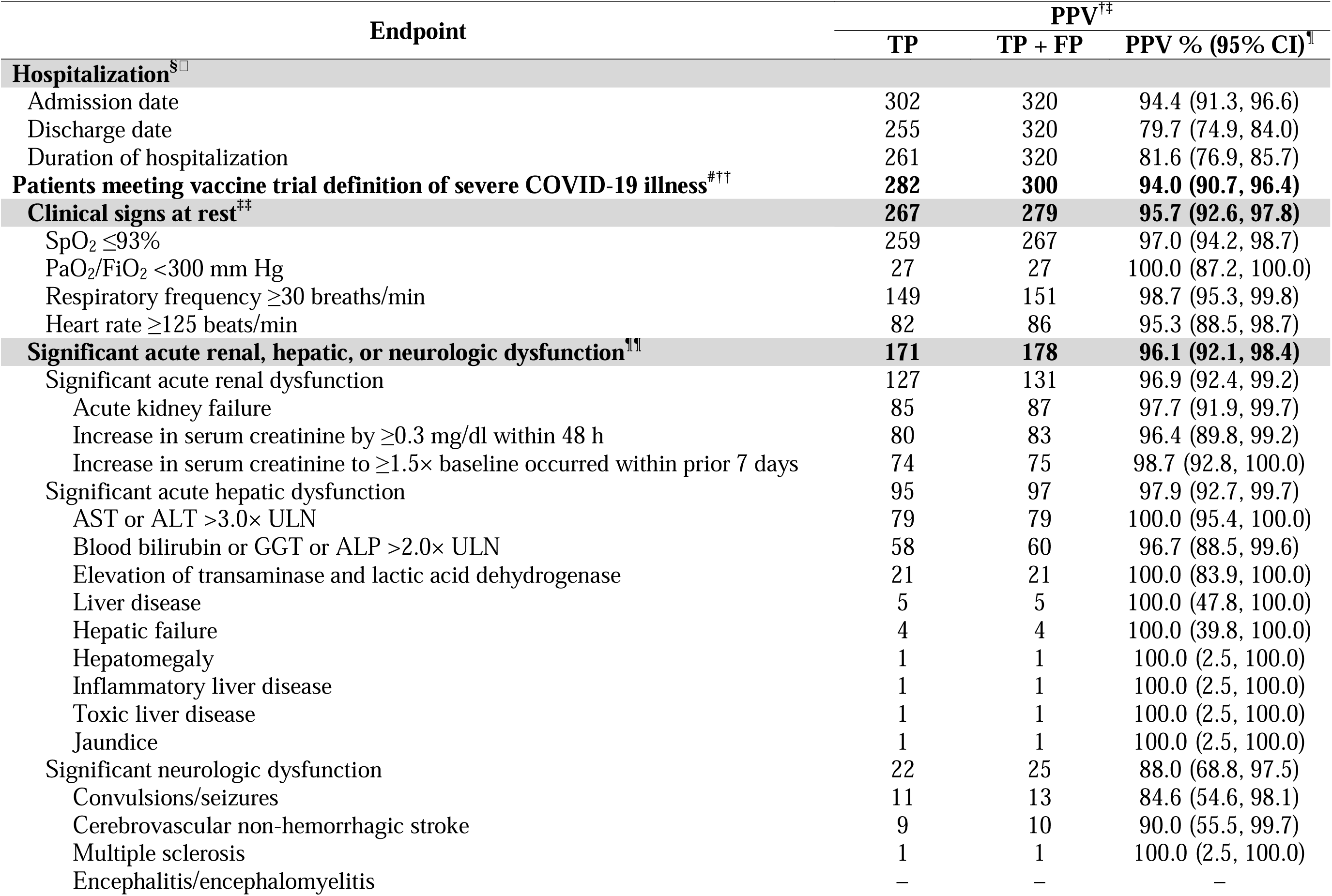
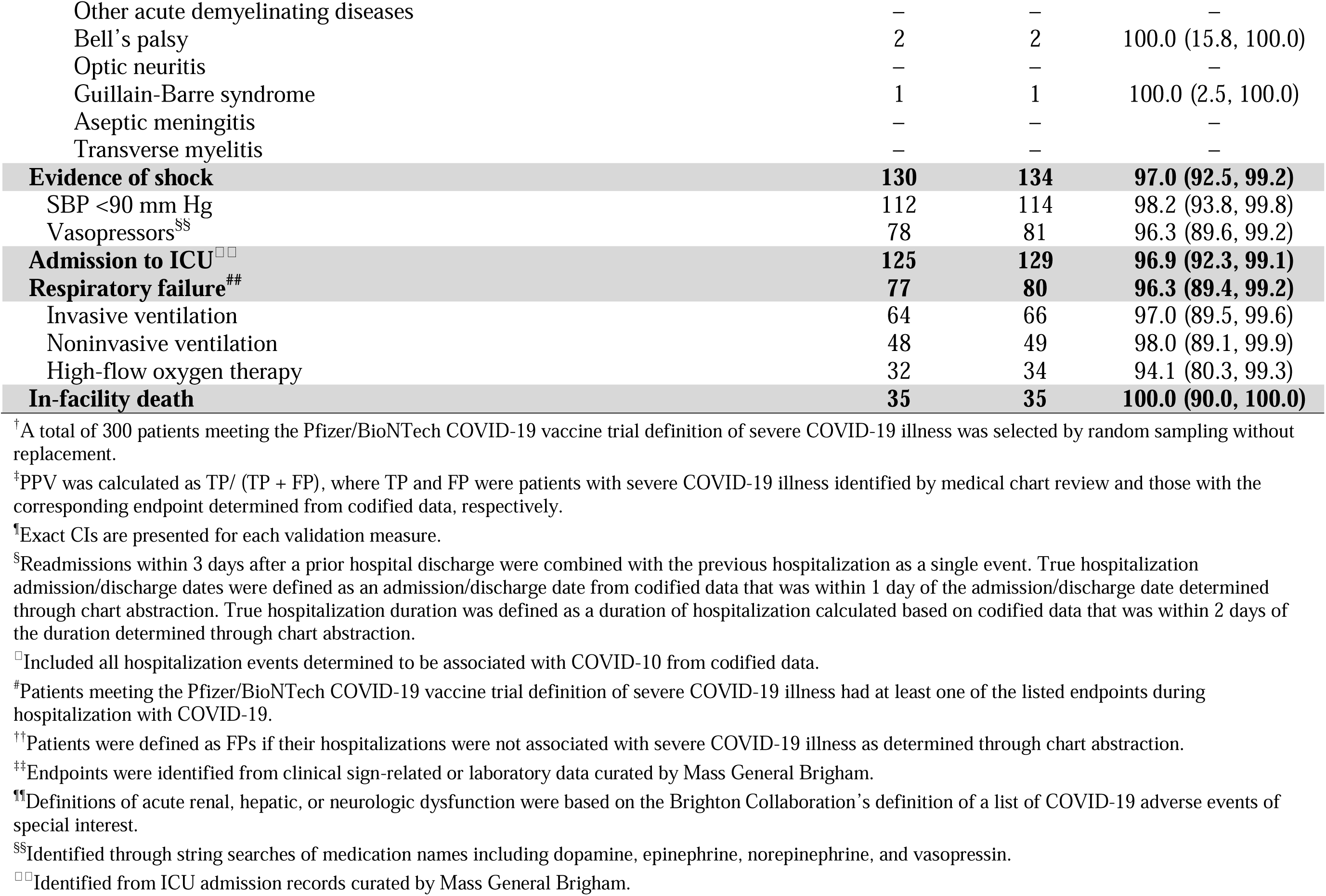
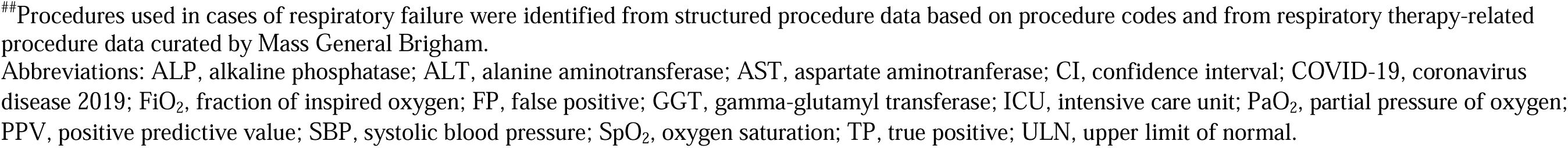
Validation of severe COVID-19 illness endpoints against medical chart review.

Severe COVID-19 illness endpoints defined by the WHO had a PPV of 78.5% in the 237 patients meeting this definition (**Table S3**). The NPV of the 100 patients who did not meet the vaccine trial definition of severe COVID-19 illness or the WHO definition of severe or critical COVID-19 illness in codified data was 75.0% and 91.7%, respectively (**Table S4**).

## DISCUSSION

Correct identification of positive cases and classification of COVID-19 illness severity are essential for assessing the efficacy and safety of vaccines and treatments in real-world settings. Observational studies are particularly important in COVID-19 research given that vaccines and treatments are initially authorized under EUA, and require supporting data on long-term efficacy and safety. Specifically, accurate COVID-19 endpoint case definitions that can be applied to observational longitudinal data are important from a methodologic standpoint. This was addressed in the present study by validating clinical trial-based COVID-19 diagnosis and severe COVID-19 illness endpoints based on the use of the ICD-10-CM code U07.1 for COVID-19 against the PCR test in a US patient cohort between March and November 2020. The results showed that the diagnosis code had low sensitivity and PPV but high specificity against the PCR test. Meanwhile, the EMR-based case definition for severe COVID-19 illness endpoints had a high PPV among patients hospitalized with COVID-19.

Following its introduction on April 1, 2020 by the Centers for Disease Control and Prevention,^16^ the ICD-10-CM code U07.1 for a confirmed diagnosis of COVID-19 was widely adopted by hospitals in the US.^17^ An analysis of inpatient hospital data from April to May 2020 in the Premier Healthcare Database reported a high sensitivity (98%) and specificity (99%) for the code compared with PCR, with a PPV of 92% and NPV of 99.97%.^18^ The lower sensitivity of the code compared with the PCR test (54%) in the present study may be explained by the inclusion in the MGB Data Mart of data from both inpatient and outpatient settings and not just hospitalized patients, who likely exhibit more severe and recognizable symptoms that facilitate accurate diagnosis. Another study that examined the concordance between coding with U07.1 and the PCR test using MGB data found that among hospitalized patients discharged between April and July 2020, the code had an overall sensitivity of 49%,^19^ which is closer to the value in the present study; however, the PPV of 90% was much higher than what we observed (23%–79%) during the same period. The impact of setting on the validity of diagnosis code U07.1 is supported by a retrospective analysis of Department of Veterans Affairs medical records from inpatient, outpatient, and emergency/urgent care settings from April 2020 to March 2021, which found that the PPV of code U07.1 was lower in outpatients settings (78% vs. 84% overall).^20^ The present study’s results suggest that the diagnosis code may have been used in cases where COVID-19– like symptoms were present but a definitive diagnosis was not confirmed by testing, or that the diagnosis code was used for billing or ordering laboratory tests. This practice may be attributable to evolving medical guidelines during COVID-19 case surges, shortage of PCR tests, or changes in hospital procedures related to PCR test administration. The reliability of code U07.1 may be improved by its combined use with the new COVID-19–related diagnosis and procedure codes released in January 2021^21^; for example, in the study by Lynch et al.,^20^ the major contributor to the FP rate for U07.1 was a history of COVID-19, for which there is now a separate code. PPV is also impacted by disease prevalence, with PPV being higher when disease prevalence is higher. As COVID-19 prevalence fluctuates over time and by geography, PPV will also vary accordingly, which may explain the variation in PPV results across studies conducted in different time periods and different populations.

The FDA recommends that COVID-19 vaccine trials evaluate severe COVID-19 illness as an endpoint.^2^ However, the definition of severe COVID-19 illness has evolved over time and there are now multiple and overlapping definitions.^2,22-24^ In the present study, 95% and 75% of patients hospitalized with COVID-19 met the BNT162b2 vaccine trial and WHO definitions of severe illness, respectively, and 41% met the WHO definition of critical illness. We assessed the validity of severe COVID-19 illness endpoints through a review of medical charts of a random sample of patients hospitalized with COVID-19. The PPV of each individual endpoint of severe illness derived from the clinical trial and included in the EMR-based case definition was >95%. Endpoints for severe COVID-19 illness according to the WHO had a comparatively lower PPV (79%), but the PPV of most WHO-defined critical COVID-19 illness endpoints was high (>91%). Thus, the BNT162b2 trial severe COVID-19 illness and WHO critical COVID-19 illness endpoints can be reliably used to identify severe cases of COVID-19 from EMR databases, which may facilitate the interpretation of COVID-19 patient data such as long-term outcomes in future studies. This is supported by recent studies that used real-world data from the US population to confirm the effectiveness of the BNT162b2 vaccine in preventing hospitalization and severe illness.^25-29^

This study had certain limitations. First data from March to November 2020 were used because the study was initiated in late 2020 and because of Institutional Review Board approval timelines, lag time for data availability, and the time required to conduct chart review, more recent data were not included. The study period covers the time prior to the availability of COVID-19 vaccines and a portion of the SARS-CoV-2 Alpha variant period. Therefore, if the patterns of use of diagnosis code U07.1 and physician recordings of COVID-19 illness signs and symptoms changed over the course of the pandemic and have been affected by vaccination status and surges of the Delta and Omicron variants, it is possible that our results cannot be extrapolated to the post-vaccination era. Second, as MGB is a regional data source centered in the Boston metropolitan area and consists of teaching hospitals affiliated with Harvard Medical School, the results may not be generalizable to all patients with COVID-19 in the US. Third, the database may have contained miscoded or incomplete information such as patient care received outside of the MGB network, which may have resulted in underreporting. Fourth, the sample sizes for the number of ICD-10-CM codes (e.g., in March or October 2020) were limited in certain analyses. Finally, validation of the EMR-based case definition for severe COVID-19 illness endpoints was based on PPV and NPV. However, PPV decreases with decreasing COVID-19 prevalence because the rate of FP increases. Thus, the fluctuation of COVID-19 prevalence could complicate the interpretation of the summary PPV estimate generated in this study as well as its generalizability to different geographic locations or time periods with surges in COVID-19 variants. Sensitivity and specificity are important measures that are not impacted by COVID-19 prevalence; however, given the logistics of chart review, their calculation would have required a large random sample of patients to ensure an adequate number of true cases, rendering sampling from true positive and negative COVID-19 severe cases infeasible.

## CONCLUSION

The ICD-10-CM diagnosis code for COVID-19 (U07.1) did not have adequate sensitivity to identify COVID-19 patients compared with the PCR test; therefore, positive cases recorded using the diagnosis code may require confirmation by PCR testing. On the other hand, the EMR-based case definition of severe COVID-19 illness endpoints showed high positive predictive performance and may be applicable to analyses of real-world COVID-19 patient datasets.

## Data Availability

All data produced in the present work are contained in the manuscript.

## ACKNOWLEDGMENTS

This study was funded by Pfizer, Inc., New York, NY, USA. The study sponsor was involved in several aspects of the research including study design, data interpretation, manuscript writing, and decision to submit the manuscript for publication. The authors thank Ann Madsen, PhD (a former employee of Pfizer) for her methodologic input in the study. Medical writing assistance was provided by Janice Imai of Analysis Group, Montreal, Canada (funding for medical writing was provided by Pfizer, Inc., New York, NY, USA).

## ETHICS STATEMENT

This study was approved by the MGB Institutional Review Board.

## PATIENT CONSENT STATEMENT

Patient consent was not required as patient data were de-identified.

