## Supplementary Figures and Tables for "Validation of Real-World Case Definitions for COVID-19 Diagnosis and Severe COVID-19 Illness Among Patients Infected with SARS-CoV-2: Translation of Clinical Trial Definitions to Real-World Settings"

### Supplementary Materials

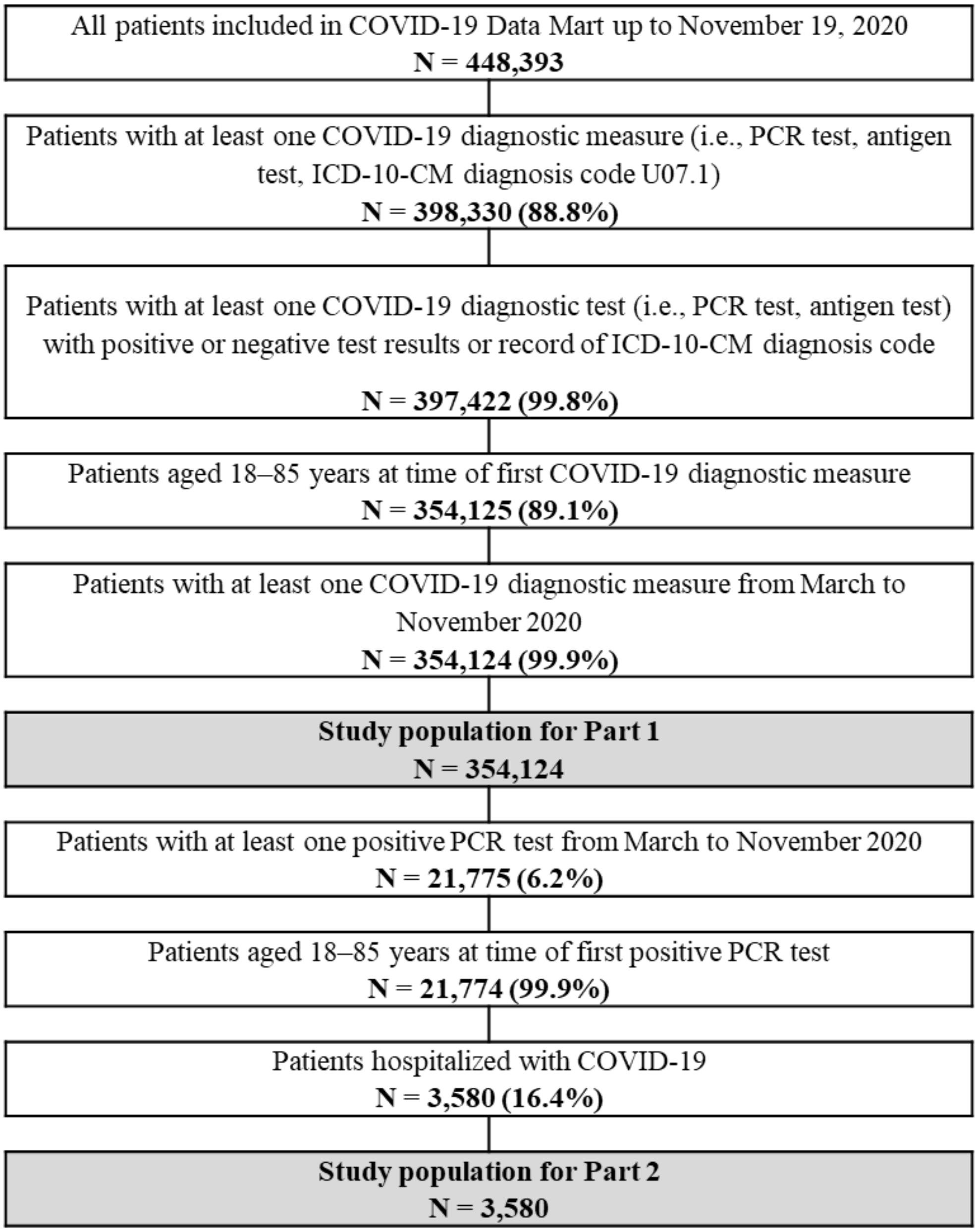


**Figure S1.** Flow diagram of patient enrollment.

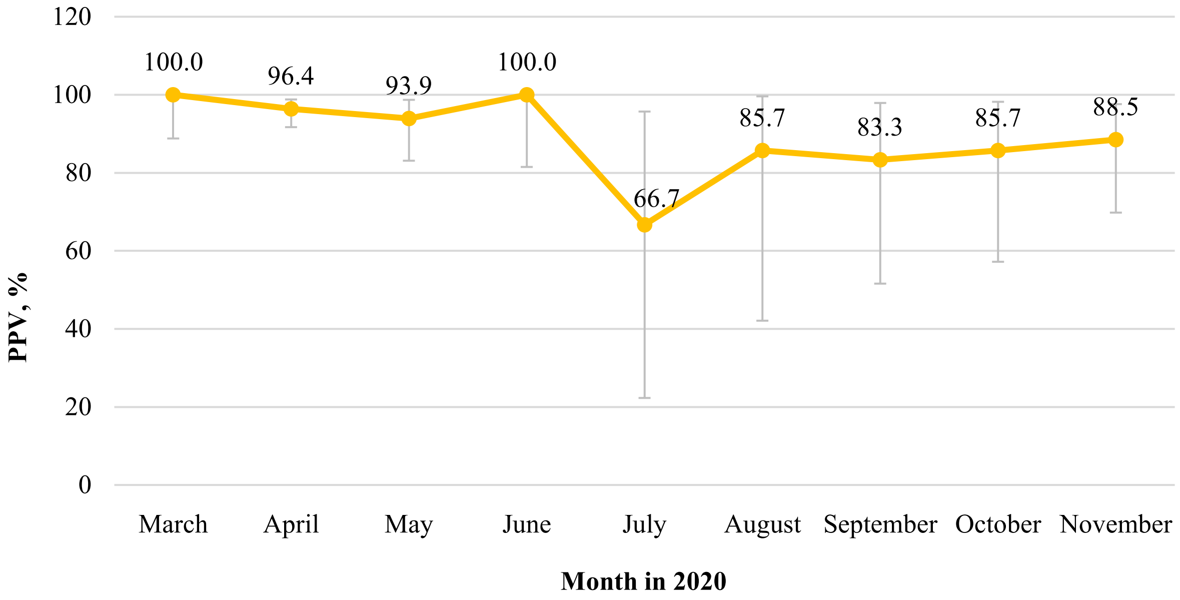


| | | | | | | | | | |
|---|---|---|---|---|---|---|---|---|---|
| **TP** | 31 | 132 | 46 | 18 | 4 | 6 | 10 | 12 | 23 |
| **TP + FP** | 31 | 137 | 49 | 18 | 6 | 7 | 12 | 14 | 26 |
| **PPV, %** | 100.0 | 96.4 | 93.9 | 100.0 | 66.7 | 85.7 | 83.3 | 85.7 | 88.5 |
| **95% CI, %** | (88.8, 100.0) | (91.7, 98.8) | (83.1, 98.7) | (81.5, 100.0) | (22.3, 95.7) | (42.1, 99.6) | (51.6, 97.9) | (57.2, 98.2) | (69.8, 97.6) |

**Figure S2.** PPV of severe COVID-19 against medical chart review by month. A total of 300 patients who met the Pfizer/BioNTech COVID-19 vaccine trial definition of severe COVID-19 illness was selected by random sampling without replacement. Data for November were available only up to November 19, 2020.

**Table S1.** COVID-19 diagnostic measures

| Diagnostic measure | Overall (March–November† 2020) N = 354,124 |
|---|---|
| **PCR** | |
| Patients with ≥1 PCR test, n (%) | 335,043 (94.6) |
| Patients with ≥1 positive test | 21,775 (6.5) |
| Patients without any positive tests | 313,268 (93.5) |
| Patients with ≥1 negative test | 323,031 (96.4) |
| Number of PCR tests per patient, mean ± SD [median] | 2.2 ± 1.7 [2.0] |
| Total number of PCR tests | 726,652 |
| Positive, n (%) | 42,930 (5.9) |
| Negative, n (%) | 683,722 (94.1) |
| **ICD-10-CM diagnosis code (U07.1)¶** | |
| Patients with ≥1 ICD-10-CM diagnosis code (U07.1), n (%) | 39,576 (11.2) |
| Number of ICD-10-CM codes per patient, mean ± SD [median] | 3.0 ± 5.4 [2.0] |

Data are shown as n (%) unless otherwise indicated.

†Data for November were available only up to November 19, 2020.

¶Defined as "COVID-19, virus identified", effective April 1, 2020.

Abbreviations: COVID-19, coronavirus disease 2019; ICD-10-CM, International Classification of Diseases 10th Revision, Clinical Modification; PCR, polymerase chain reaction; SD, standard deviation.

**Table S2.** Frequency of severe or critical COVID-19 illness endpoints per WHO definition in patients hospitalized with COVID-19

| Endpoint | All patients hospitalized with COVID-19 N = 3,580 |
|---|---|
| **Patients meeting WHO definition of severe or critical COVID-19 illness, n (%)** | **2,822 (78.8)** |
| **Patients meeting WHO definition of severe COVID-19 illness[†], n (%)** | **2,697 (75.3)** |
| $SpO_2$[‡] | |
| Patients with $SpO_2$ available | 3,578 (99.9) |
| $SpO_2$ <90% | 1,931 (54.0) |
| Respiratory frequency[‡] | |
| Patients with respiratory frequency available | 3,573 (99.8) |
| Respiratory frequency >30 breaths/min | 1,626 (45.5) |
| Signs of severe respiratory distress[¶] | 1,705 (47.6) |
| **Patients meeting WHO definition of critical COVID-19 illness[§], n (%)** | **1,454 (40.6)** |
| Requiring life-sustaining treatment | 994 (27.8) |
| Vasopressors[‖] | 914 (25.5) |
| Invasive ventilation[#] | 738 (20.6) |
| Noninvasive ventilation[#] | 605 (16.9) |
| Acute respiratory distress syndrome[¶] | 1,167 (32.6) |
| Sepsis[¶] | 494 (13.8) |
| Septic shock[¶] | 334 (9.3) |

[†]Patients meeting the WHO definition of severe COVID-19 illness had at least one of the listed endpoints during hospitalization with COVID-19.

[‡]If a patient had multiple measurements on the same day, the most severe result was used.

[¶]Identified using diagnosis codes.

[§]Patients meeting the WHO definition of critical COVID-19 illness had at least one of the listed endpoints during hospitalization with COVID-19.

[‖]Identified through string searches of medication names including dopamine, dobutamine, milrinone, levosimendan, epinephrine, norepinephrine, vasopressin, and phenylephrine.

[#]Identified from structured procedure data based on procedure codes and from respiratory therapy-related procedure data curated by Mass General Brigham.

Abbreviations: COVID-19, coronavirus disease 2019; $SpO_2$, oxygen saturation; WHO, World Health Organization.

**Table S3.** Validation of severe or critical COVID-19 illness endpoints per WHO definition against medical chart review

| **Endpoint** | **PPV[†‡]** | | |
|---|---|---|---|
| | **TP** | **TP + FP** | **PPV % (95% CI)[¶]** |
| **Patients meeting WHO definition of severe or critical COVID-19 illness** | **190** | **244** | **77.9 (72.1, 82.9)** |
| **Patients meeting WHO definition of severe COVID-19 illness[§]** | **186** | **237** | **78.5 (72.7, 83.5)** |
| $SpO_2$ <90%[‖] | 157 | 172 | 91.3 (86.0, 95.0) |
| Respiratory frequency >30 breaths/min | 127 | 136 | 93.4 (87.8, 96.9) |
| Signs of severe respiratory distress[#] | 111 | 149 | 74.5 (66.7, 81.3) |
| **Patients meeting WHO definition of critical COVID-19 illness[††]** | **108** | **119** | **90.8 (84.1, 95.3)** |
| Requiring life-sustaining treatment | 79 | 85 | 92.9 (85.3, 97.4) |
| Vasopressors[‡‡] | 76 | 81 | 93.8 (86.2, 98.0) |
| Invasive ventilation[¶¶] | 64 | 66 | 97.0 (89.5, 99.6) |
| Noninvasive ventilation[¶¶] | 48 | 49 | 98.0 (89.1, 99.9) |
| Acute respiratory distress syndrome[#] | 95 | 102 | 93.1 (86.4, 97.2) |
| Sepsis[#] | 51 | 52 | 98.1 (89.7, 100.0) |
| Septic shock[#] | 37 | 38 | 97.4 (86.2, 99.9) |

[†]Validation was conducted in a subset of the 300 patients who met the WHO definition of severe or critical COVID-19 illness based on codified data.

[‡]PPV was calculated as TP/ (TP + FP), where TP and FP were patients meeting the WHO definition of severe COVID-19 illness by medical chart review and those with the corresponding endpoint determined from codified data, respectively.

[¶]Exact CIs are presented for each validation measure.

[§]Patients who met the WHO definition of severe COVID-19 illness had at least one of the listed endpoints during hospitalization with COVID-19.

[‖]Endpoints were identified from clinical sign-related or laboratory data curated by Mass General Brigham.

[#]Identified from structured diagnosis data based on diagnosis codes.

[††]Patients who met the WHO definition of critical COVID-19 illness had at least one of the listed endpoints during hospitalization with COVID-19.

[‡‡]Identified through string searches of medication names including dopamine, epinephrine, norepinephrine, and vasopressin.

[¶¶]Identified from structured procedure data based on procedure codes and from respiratory therapy-related procedure data curated by Mass General Brigham.

Abbreviations: CI, confidence interval; COVID-19, coronavirus disease 2019; FP, false positive; PPV, positive predictive value; $SpO_2$, oxygen saturation; TP, true positive; WHO, World Health Organization.

**Table S4.** Validation of non-severe COVID-19 illness in codified data against medical chart review

| Endpoint | NPV[†‡] | | |
|---|---|---|---|
| | TN | TN + FN | NPV % (95% CI)[¶] |
| Patients not meeting vaccine trial definition of severe COVID-19 illness[§] | 75 | 100 | 75.0 (65.3, 83.1) |
| Patients not meeting WHO definition of severe or critical COVID-19 illness | 66 | 72 | 91.7 (82.7, 96.9) |

[†]NPV was calculated based on a subgroup of 100 patients who did not meet the Pfizer/BioNTech COVID-19 vaccine trial definition of severe COVID-19 illness based on codified data.

[‡]NPV was calculated as TN/ (TN + FN), where TN and FN were patients who did not meet the definition of severe COVID-19 illness based on chart review and codified data, respectively.

[¶]Exact CIs are presented for each validation measure.

[§]Patients with hospitalizations that were determined to not be COVID-19–related through chart review were categorized as TN.

Abbreviations: CI, confidence interval; COVID-19, coronavirus disease 2019; FN, false negative; NPV, negative predictive value; TN, true negative; WHO, World Health Organization.
